# Accentuated Divergence, Constrained Expression: Autistic Traits Modulate the Creative Brain in Adults with ADHD

**DOI:** 10.64898/2026.10.20.26365994

**Authors:** Ningning Liu, Zelin Mu, Xueyang Wang, Gaoding Jia, Alitanshunbuer, Jiabin Liu, Jinhua Xu, Haimei Li, Yufeng Wang, Jiang Qiu, Qiujin Qian, Lu Liu

## Abstract

**Background:** Attention-deficit/hyperactivity disorder (ADHD) is a heterogeneous neurodevelopmental condition, and co-occurring autistic traits may shape its phenotypic expression. Although creativity has been discussed as a relative strength in ADHD, it remains unclear whether autistic traits are associated with different dimensions of creativity in adults with ADHD and whether large-scale brain networks contribute to these associations.

**Methods:** We compared divergent thinking (DT) and real-life creativity between 65 adults with ADHD and 73 neurotypical controls. We used subgroup comparisons, partial correlation analyses, and multiple regression models to determine the effects of autistic traits (ATs) on creativity dimensions within the ADHD cohort. Guided by prior creativity-network research, we analyzed resting-state functional connectivity (FC) of the Default Mode Network (DMN) and Frontoparietal Network (FPN) to uncover the neural mechanisms.

**Results:** The ADHD cohort demonstrated significantly higher scores on both DT (*p*s = 0.002–0.009) and real-life creativity measures (*p*s = 0.011–0.017) compared to the neurotypical control group. Importantly, increased ATs within the ADHD group were associated with a dissociated creativity profile, showing positive associations with DT but negative associations with real-life creativity. At the neural level, stronger right DMN–FPN connectivity mediated positive associations between ATs and DT performance (ps = 0.007–0.018), but mediated a negative association between ATs and self-reported imagination (p = 0.029).

**Conclusions:** In individuals with ADHD, co-occurring ATs exert a paradoxical effect on creativity, enhancing DT while compromising real-life creative achievement. In this subgroup with ADHD and elevated ATs, interventions should capitalize on ATs-associated ideational strengths while circumventing translational barriers, to enhance real-world creative outcomes.

## 1. Introduction

Attention-deficit/hyperactivity disorder (ADHD) has traditionally been regarded as a neurodevelopmental condition with childhood onset, characterized by challenges in behavior, cognitive function, and various other domains [1]. Prior research has largely focused on the deficits associated with ADHD and emphasized the need for external interventions to alleviate their symptoms [2]. Recently, the neurodiversity paradigm has received increasing attention which posits that neurodivergent conditions such as ADHD should be regarded as variations rather than deficiencies, impairments, or disorders [3, 4]. Emphasizing the strengths of neurodivergent individuals can help them flourish in specific contexts, with creativity being one of the most frequently discussed attributes [4, 5].

Creativity is usually regarded as the ability to produce ideas or products that are both original and useful [6, 7]. Creative thinking is a core cognitive process that researchers focus on, it is generally considered to comprise two cognitive components: divergent thinking (DT) and convergent thinking (CT) [8]. DT emphasizes a person’s ability to generate multiple and diverse ideas in response to open-ended problems, which can be assessed using various methods, among which the Alternate Uses Task (AUT) is currently the most widely adopted [5]. CT refers to the ability to integrate information and narrow down multiple possibilities to the best solution [8]. The widely used test is the Remote Associates Task (RAT) [9]. Another widely used dimension for assessing creativity is real-life creativity. One example is the Creative Achievement Questionnaire (CAQ), which can be used to assess an individual’s past creative accomplishments across various domains [10]. For individuals with ADHD, some empirical studies have reported enhanced performance in DT [11]. In addition, several studies have suggested a positive association between ADHD and real-life creativity, indicating that ADHD may be more likely to attain real-world accomplishments in creative domains [12]. Still, current results are not entirely concordant. Given the large heterogeneity of ADHD, many factors, including co-occurring autistic traits, may contribute to these inconsistent findings. However, limited related studies have been conducted on this topic.

Autism, which commonly co-occurs with ADHD, is traditionally defined by persistent difficulties in social interaction and communication, along with restricted, repetitive patterns of behavior or interests [1, 13]. Prior studies have reported that approximately 10%∼20% of individuals with ADHD exhibit co-occuring autism or elevated autistic traits (ATs) [13–16]. Interestingly, autistic individuals or individuals with elevated ATs have also been reported to show unique creative potential [17, 18]. However, autism-creativity relationships seem to be less consistent than that between ADHD symptoms and creativity [11]. Some evidence even suggests that the creativity advantages observed in autistic individuals may be partly due to co-occurring ADHD symptoms [19]. Other work suggests that the impact of autism on creativity may be dimension-specific [20]. Individuals with autism tend to exhibit lower levels of DT performance in fluency and flexibility, but may demonstrate a high degree of originality in DT [20]. These findings raise the possibility that co-occurring ATs may relate to ADHD-related creativity in complex and dimension-specific ways. However, previous studies often treated ADHD-driven creativity in isolation, leaving the role of co-occurring ATs underexplored. Furthermore, although creativity has been studied for over a century and a substantial body of neuroimaging research on creativity has been conducted in neurotypical populations [21, 22], neural studies focusing on creativity in individuals with ADHD remain limited [5]. Research in general neurotypical populations suggests that creative thinking may involve functional connectivity (FC) across multiple networks, particularly the frontoparietal network (FPN) and the default mode network (DMN) [23]. It remains unknown whether the potential mechanism through which ATs influences the creativity of individuals with ADHD can be explained by these neural structures.

To address these gaps, the present study employed a sample of 65 adults who strictly met the diagnostic criteria for ADHD, alongside 73 control participants. We systematically integrated assessments of creativity—encompassing DT, CT, and real-life measures—alongside evaluations of brain functional network connectivity. This approach aimed to provide a more comprehensive understanding of the relationship between ATs and creativity within the ADHD cohort, as well as to further investigate the underlying neural mechanisms associated with these differences.

## 2. Methods

The overall design and analytic procedure of the study is illustrated in **Figure 1**. The present study was approved by the local institutional review board (Approval Number: 2024-Ethics Rev.16).

**Figure 1.**
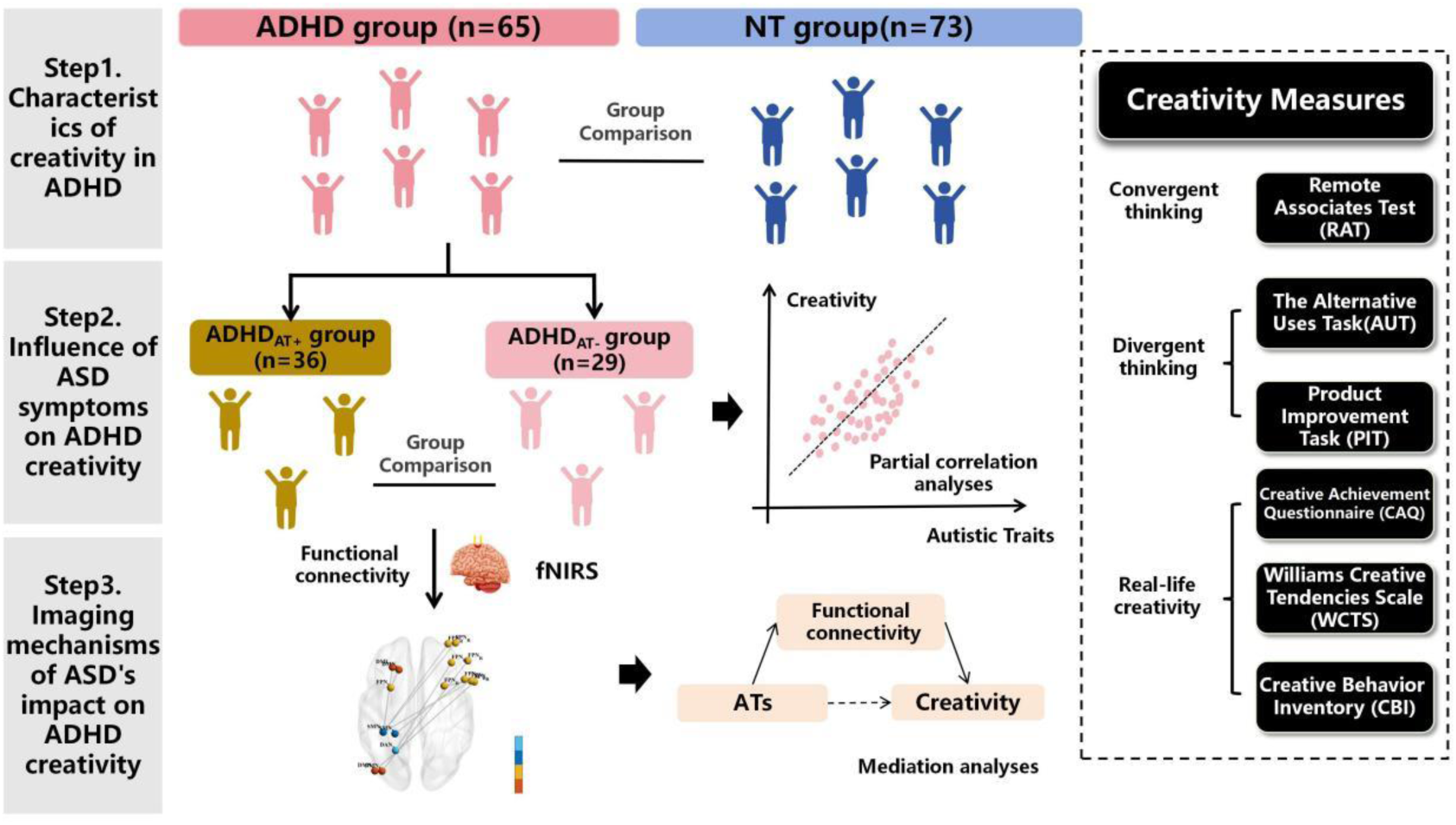
Flowchart of the study procedure. Note: The study included 65 adults with ADHD and 73 neurotypical controls. Step 1: Group comparisons were conducted to assess characteristics of creativity in ADHD. Step 2: ADHD participants were divided into high (ADHD_AT+_, n = 36) and low (ADHD_AT-_, n = 29) autistic-trait subgroups for further comparison and partial correlation analyses. Step 3: Resting-state fNIRS was used to examine functional connectivity, followed by correlation analyses and mediation analyses. Creativity measures included convergent thinking, divergent thinking, and real-life creativity.

### 2.1 Participants

We recruited a total of 138 adults, including 65 diagnosed with ADHD and 73 controls in this study. Experienced psychiatrists interviewed all ADHD participants according to the criteria of the Diagnostic and Statistical Manual of Mental Disorders, Fifth Edition (DSM-5) [1], with reference to the Conners’ Adult ADHD Diagnostic Interview [24]. All included ADHD participants in this study were (1) aged 18-45 years; (2) had no history of severe physical disease; and (3) had no history of schizophrenia, clinically significant panic disorders. The controls were from the community population who: (1) aged 18-45 years; (2) no current or past psychiatric disorders and no family history of psychiatric illness; and (3) never having used psychotropic medication.

### 2.2 Clinical measures

#### ADHD symptoms

We assessed ADHD symptoms using ADHD Rating Scale-IV (ADHD-RS-IV) [25]. This questionnaire is a widely - used tool, which consists of the inattention dimension (the first 9 items) and the hyperactivity/impulsivity dimension (the last 9 items). Each item is rated on a Likert-type scale range from 0 (not at all) to 3 (very often), based on the symptom frequency. We calculated total scores as the sum of both dimensions.

#### Autistic traits

To assess autistic traits, the participants filled out the Autism-Spectrum Quotient (AQ) [26]. This 50-item questionnaire asks participants to indicate agreement with statements on a scale from 1 (definitely agree) to 4 (definitely disagree). Responses were scored according to the standard AQ scoring procedure, yielding a total score from 0 to 50. A greater score indexes more pronounced autistic traits. The AQ questionnaire has a high discriminant validity and satisfactory to good internal consistency [27]. We divided adult ADHD into high autistic traits group (ADHD_AT+_) and low autistic traits group (ADHD_AT-_) with a 23-point boundary for AQ scores, which is believed to be able to identify people who are most likely to have a clinical level of ATs [28, 29].

### 2.3 Creative assessment

#### 2.3.1 Divergent Thinking

Alternate Uses Task (AUT) and Product Improvement Task (PIT) were utilized to evaluate DT. AUT emphasizes spontaneous, unconventional idea generation, while PIT, adapted from Torrance’s creativity measures, prompts participants to propose goal-directed enhancements to familiar items. Together, they provide a broader evaluation of DT [30].

##### AUT

Participants were asked to generate as many novel and original ways as possible for a “brick” or “can” in 3 min. For each item, three trained coders counted all ideas independently to determine fluency and coded all ideas to determine flexibility and originality. Fleiss’ Kappa coefficients for fluency, flexibility, and originality were 0.82, 0.76, and 0.79, respectively. In the present study, all three dimensions were included as indicators of DT [31], and the total score was computed as the sum of the standardized fluency, flexibility, and originality scores.

##### PIT

PIT requires participants to list the most ingenious, interesting, and unusual ways to make a toy elephant more fun, enjoyable and appealing in a 10-min time period [30, 32]. Responses were also scored on fluency, flexibility and originality of PIT. Fleiss’ Kappa coefficients for fluency, flexibility, and originality were 0.75, 0.58, and 0.68, respectively. In the present study, we used both the three dimensions (fluency, flexibility, and originality) as well as their composite mean score for subsequent analyses.

Supplementary Material provides detailed scoring processes for the AUT and PIT tasks.

#### 2.3.2 Convergent Thinking

The Remote Associates Test (RAT) was used to assess CT. This task consists of 30 items, in which participants were asked to come up with one word that connects all three given words. For example, one of the items contains the words “拍(pāi)”, “买(mǎi)”, “贩(fàn)” where the correct answer is “卖(mài)”. The total scores were calculated as the number of correctly solved items, with a range of 0 to 30. The RAT has been found to have good internal consistency (Cronbach’s alpha = 0.85) and convergent validity with other convergent thinking tasks (r = 0.32-0.39) [9].

#### 2.3.3 Real-life creativity

To comprehensively assess creativity from multiple dimensions, we employed three widely used instruments to evaluate real-life creativity: the CAQ, the Williams Creative Tendencies Scale (WCTS), and the Creative Behavior Inventory (CBI). The CAQ is designed to quantify domain-specific creative accomplishments across ten distinct fields [10]. The WCTS assesses trait level proclivities that predispose individuals toward creative thought and action [33]. The CBI measures the frequency of everyday creative behaviors by recording how often participants engage in various creative activities in their daily lives [34].

##### CAQ

The CAQ is a self-report questionnaire that assesses creative achievement across ten domains: visual arts, music, dance, architectural design, creative writing, humor, inventions, scientific discovery, theatre and film, and culinary arts. Each of the ten domains has eight ranked questions weighted with a score of 0-7. Participants were asked to indicate how many times they earned each achievement. Total scores were computed by adding up all scales. The Chinese version of the CAQ showed substantial test-retest reliability, with a test-retest reliability coefficient of 0.77 [35].

##### WCTS

The WCTS is composed of 50 items, and the subjects were asked to respond to a 3-point Likert-type scale. WCTS includes four subscales, namely, curiosity, imagination, challenge and adventurousness [33]. Reliability analysis showed that the test-retest reliability coefficient was 0.702, Cronbach’s alpha coefficient was about 0.8 [33].

##### CBI

The inventory is a widely used measure of everyday creativity, which includes 28 items in the literary, performing and visual arts, and crafts. The questionnaire requires subjects to indicate the number of creative activities they have performed on a 4-point scale. A total score was derived by adding all scores together. Recent psychometric evidence supports the reliability with Cronbach’s alpha reported as 0.91 [34].

Supplementary Material provides detailed scoring processes and additional information for the CAQ, WCTS, and CBI tasks.

### 2.4 fNIRS Data Acquisition and Processing

#### fNIRS Data Acquisition

Previous resting-state neuroimaging studies have linked individual differences in creativity to large-scale functional connectivity patterns, particularly involving cortical networks such as the DMN and FPN [36, 37], making fNIRS an ideal modality with which to study creativity. Quantitative studies have demonstrated the feasibility and utility in characterizing brain activation and functional connectivity [38, 39].

In this study, a 24-source/28-detector multichannel fNIRS system (Nirscan, Hui Chuang, China) was configured to generate 80 measurement channels with a fixed source-detector distance of 3 cm covering the frontal, occipital and parietal lobes. For each subject, approximately 12 minutes of resting-state data were acquired at a sampling rate of 17 Hz. Environmental conditions were kept stable throughout the session [40]. The method for obtaining the Montreal Neurological Institute (MNI) coordinates and the corresponding anatomical positions of each channel is available in a previous study [40].

#### Data preprocessing

FC-NIRS package was used to preprocess the collected fNIRS data [41]. The preprocessing steps included channel exclusion, signal conversion, motion artifact correction, band-pass filtering, hemoglobin concentration calculation, and dataset truncation (see Supplementary Materials for details).

#### Calculation of functional connectivity

Based on the creativity-related cortical networks described above, we focused on FC within and between the DMN and FPN in this study. FC was calculated using the FC-NIRS toolbox [41]. Specifically, an 80×80 correlation matrix was generated for each participant by calculating the Pearson correlation coefficients between the time series of all possible pairs of fNIRS channels. Following previous fNIRS procedures [40], fNIRS channel coordinates were projected onto a standardized cortical surface and assigned to large-scale brain networks using Yeo et al.’s network template [42]. Based on this network assignment, we averaged channel-level FC values within and between the bilateral DMN and FPN to obtain ten candidate DMN/FPN-related FC indices for each participant, including four within-network indices and six between-network indices. These correlation coefficients were then Fisher z-transformed to improve normality for subsequent group-level analyses.

Details regarding the acquisition of fNIRS data, preprocessing and FC calculation are available in the Supplementary Material.

### 2.5 Statistical Analysis

#### 2.5.1 Characteristics of creativity in ADHD

To examine overall group differences between adults with ADHD and controls, two-sample t-tests were used in creative task outcomes for continuous, normally-distributed data. Mann-Whitney U tests were conducted for non-normally distributed data. In addition, partial correlation analyses were performed to investigate how different dimensions of ADHD symptoms contribute to creativity.

#### 2.5.2 Influence of ATs on ADHD creativity

Within the ADHD group, we examined the influence of ATs on creativity from both categorical and dimensional perspectives. First, the creativity scores of ADHD_AT-_ and ADHD_AT+_ were compared using two-sample t-tests or Mann-Whitney U test. Then, partial correlation analyses were performed to explore the associations between autistic, ADHD symptom severity and creativity outcomes with age, sex and IQ controlled as covariates. In addition, we conducted multiple linear regression analyses as a complementary approach to the partial correlation, with creativity measures as dependent variables and AQ scores and ADHD symptom scores as main predictors, while age, sex, and IQ were included as covariates.

#### 2.5.3 Imaging mechanisms of ATs impact on ADHD creativity

Resting-state fNIRS was used to explore neural mechanisms. To further examine whether the FCs between large-scale brain networks mediates the association between ATs and creativity in adults with ADHD, mediation analyses were conducted.

Prior to mediation, correlation analyses using listwise exclusion were performed within the ADHD group to ensure consistent case inclusion across variables. Only FC indices that showed associations with both ATs and creativity measures were entered as candidate mediators. In each model, the total score of the AQ was entered as the independent variable (X), creativity indices were entered as dependent variables (Y), and FC indices between key brain networks (e.g., DMN– FPN) were entered as mediators (M). Age, sex, and IQ were included as covariates in both the mediator model predicting FC from AQ scores and the outcome model predicting creativity from AQ scores and FC. Indirect effects were estimated using bias-corrected bootstrapping with 5,000 resamples, and 95% percentile confidence intervals were employed to assess statistical significance. An effect was deemed significant if the confidence interval did not include zero.

All statistical analyses were conducted using IBM SPSS Statistics (version 27). fNIRS-related analyses were performed using MATLAB (R2021a) and BrainNet Viewer (Version 1.7). Unless otherwise specified, a two-tailed *p* < 0.05 was considered statistically significant. For analyses involving multiple comparisons, false discovery rate (FDR) correction was applied, and results surviving *q* (FDR-corrected) < 0.05 were reported in the text.

## 3. Results

### 3.1 Characteristics of creativity in ADHD

A cohort comprising 65 adults diagnosed with ADHD and 73 controls was recruited. No significant differences in age or sex distribution were observed between groups (**Table 1**).

**Table 1.**
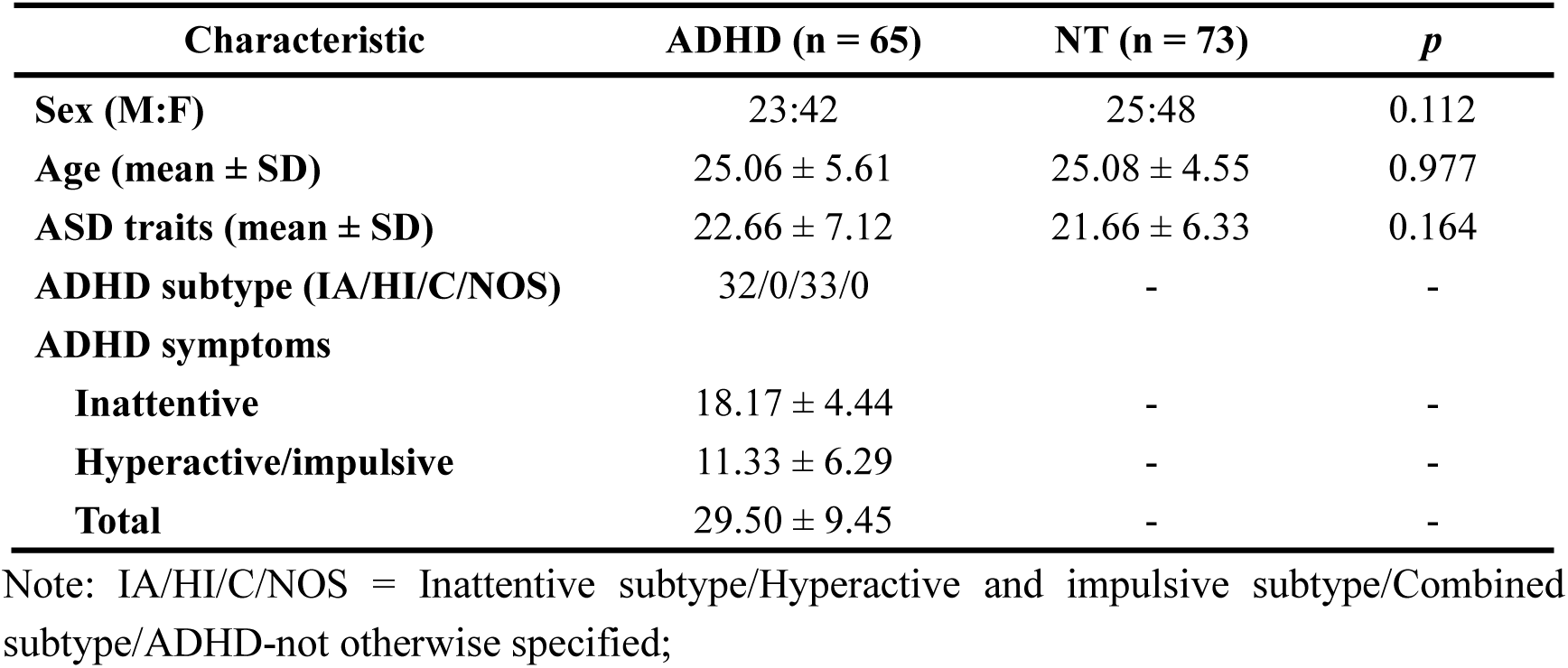
Demographic and clinical variables of adult ADHD and NT.

| Characteristic | ADHD (n = 65) | NT (n = 73) | *p* |
| --- | --- | --- | --- |
| **Sex (M:F)** | 23:42 | 25:48 | 0.112 |
| **Age (mean ± SD)** | 25.06 ± 5.61 | 25.08 ± 4.55 | 0.977 |
| **ASD traits (mean ± SD)** | 22.66 ± 7.12 | 21.66 ± 6.33 | 0.164 |
| **ADHD subtype (IA/HI/C/NOS)** | 32/0/33/0 | - | - |
| **ADHD symptoms** |  |  |  |
| **Inattentive** | 18.17 ± 4.44 | - | - |
| **Hyperactive/impulsive** | 11.33 ± 6.29 | - | - |
| **Total** | 29.50 ± 9.45 | - | - |

Note: IA/HI/C/NOS = Inattentive subtype/Hyperactive and impulsive subtype/Combined subtype/ADHD-not otherwise specified;

Compared to controls, individuals with ADHD demonstrated enhanced DT and real-life creativity. Specifically, in the PIT, ADHD participants exhibited significantly greater originality [(2.95 ± 1.04) vs.(2.21 ± 1.10), *p =* 0.002], fluency [(11.22 ± 6.03) vs. (8.13 ± 7.49), *p* = 0.007], flexibility [(10.11 ± 5.30) vs. (7.29 ± 6.52), *p* = 0.009] and average score [(8.09 ± 4.96) vs.(5.88 ± 5.00), *p* = 0.007]. Similarly, in the WCTS, ADHD scored higher than the controls on the total score[(111.40 ± 15.00) vs.(106.79 ± 12.04), *p* = 0.011], as well as on the “Challenge” [(28.43 ± 3.64) vs.(27.09 ± 3.17), *p* = 0.017] and “Imagination” subscales [(27.67 ± 4.69) vs.(25.48 ± 4.47), *p* = 0.011]. However, no significant group differences were found in CT (**Figure 2A**).

**Figure 2.**
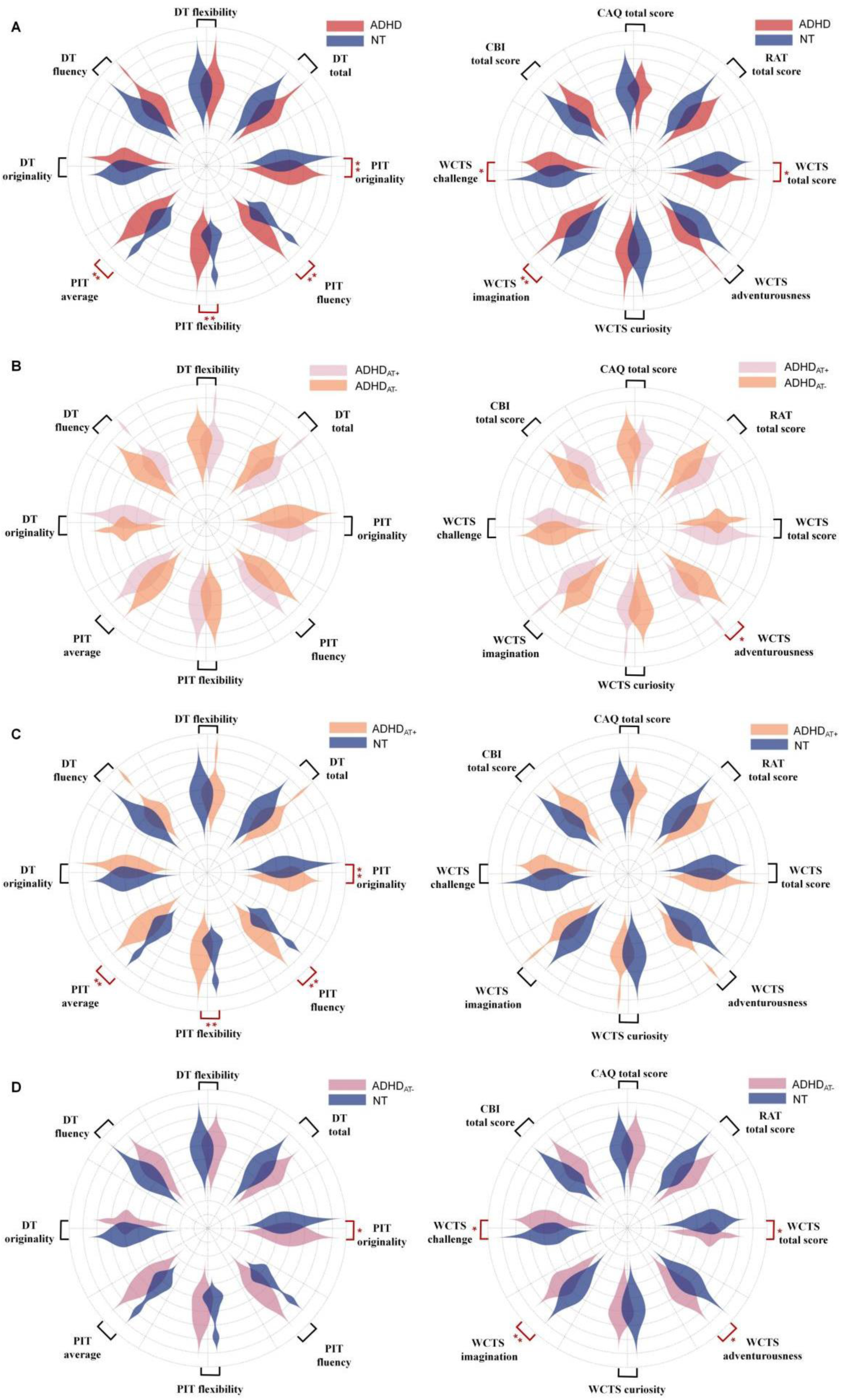
Group comparisons of creativity measures between ADHD, ADHD subgroups, and control. Note: (A) Creativity profiles of ADHD vs. NT. (B) Creativity profiles of ADHD_AT+_ vs. ADHD_AT-_. (C) Creativity profiles of ADHD_AT+_ vs. NT. (D) Creativity profiles of ADHD_AT-_ vs. NT. AUT: The Alternate Uses Task; PIT: Product Improvement Task; WCTS: Williams Creative Tendencies Scale; CBI: Creative Behavior Inventory; CAQ: Creative Achievement Questionnaire; PIT: Product Improvement Task. *p<0.05; **p<0.01. Note: All creativity scores were z-transformed for visualization purposes to enable comparison across tasks with different scoring scales.

The correlation analysis showed that hyperactivity/impulsivity symptoms exhibited significant positive correlations with the total score of the WCTS (*r* = 0.325, *p* = 0.019), as well as its “Adventurousness” (*r* = 0.304, *p* = 0.029) and “Challenge” subscales (*r* = 0.391, *p* = 0.004). In contrast, no significant associations were observed between inattention symptoms and any creativity metrics (see **Supplementary Figure S1**).

To further clarify these associations, we conducted an exploratory correlation analysis by separately including the hyperactivity (HY) and impulsivity (IMP) subscores. Results revealed that IMP, rather than HY, showed stronger positive associations with the WCTS total score (*r* = 0.392, *p* = 0.004) and its “Adventurousness” (*r* = 0.333, *p* = 0.017), “Curiosity” (*r* = 0.295, *p* = 0.035), “Challenge” (*r* = 0.389, *p* = 0.005) and “Imagination” (*r* = 0.373, *p* = 0.007) subscales, suggesting that the overall effect of hyperactivity/impulsivity on creativity may be primarily driven by impulsivity symptoms.

### 3.2 Influence of ATs on ADHD creativity

Based on the AQ scores, we divided the ADHD group into ADHD_AT+_ group (n = 36) and ADHD_AT-_ group (n = 29). These two subgroups were well-matched in terms of gender ratio (ADHD_AT+_: 13M/23F; ADHD_AT-_: 10M/19F), age (25.14 ± 5.96 vs. 24.97 ± 5.53), and intelligence quotient (122.38 ± 8.97 vs. 121.65 ± 11.21) (all *p*-values > 0.05). As expected, the ADHD_AT+_ group showed significantly higher autistic trait levels than the ADHD_AT-_ group (28.36 ± 4.34 vs. 16.31 ± 3.52, *p* < 0.001).

When directly comparing ADHD_AT+_ and ADHD_AT-_ subgroups, the ADHD_AT+_ group only scored higher on the WCTS adventurousness dimension (*p* = 0.013) (**Figure 2B**). For better clarity, we also present comparisons between the different ADHD groups and the NT. When comparing with NT, the ADHD_AT+_ group continued to exhibit significantly better DT, with significantly higher scores in PIT indices (originality, fluency, flexibility, and total scores; all *p* < 0.01). However, no significant group differences were observed for real-life creativity measures (**Figure 2C**). In contrast, the ADHD_AT-_ group exhibited significantly higher scores in real-life creativity, as reflected by elevated WCTS dimensions (including Imagination, Challenge, and Total scores; all *p* < 0.05), while the significance in DT was largely diminished, with only the “originality” component in the PIT remaining significant (*p* = 0.048) (**Figure 2D**).

Partial correlation analysis (controlling for age, sex, IQ) revealed that AQ scores did not demonstrate a significant association with any measure of creativity. Notably, although not significantly, AQ scores showed predominantly negative associations with WCTS dimensions. Conversely, higher ADHD symptom scores were positively correlated with the “Challenge” dimension of the WCTS (see Supplementary **Figure S2**).

To further validate the above results, we employed multiple regression analysis (see Supplementary **Table S1**). Consistent with previous results, ADHD symptoms were positive predictors of the “Challenge” dimension in the WCTS (*β* = 0.119, *p* = 0.014). AQ coefficients were negative across all WCTS subscales, but none reached statistical significance (standardized *β* ranged from −0.22 to −0.04, all *p* > 0.05).

### 3.3 Imaging mechanisms of ATs impact on ADHD creativity

The arrangement of the 80 fNIRS measurement channels is presented in **Figure 3A**. We performed an additional correlation analysis using listwise exclusion within the ADHD group. At the nominal p < 0.05 level, this analysis revealed that FC between right DMN and right FPN (R_DMN-R_FPN) was significantly associated with ATs as well as PIT and WCTS indices. Accordingly, R_DMN-R_FPN was selected as the candidate mediator (**Figure 3B**).

**Figure 3.**
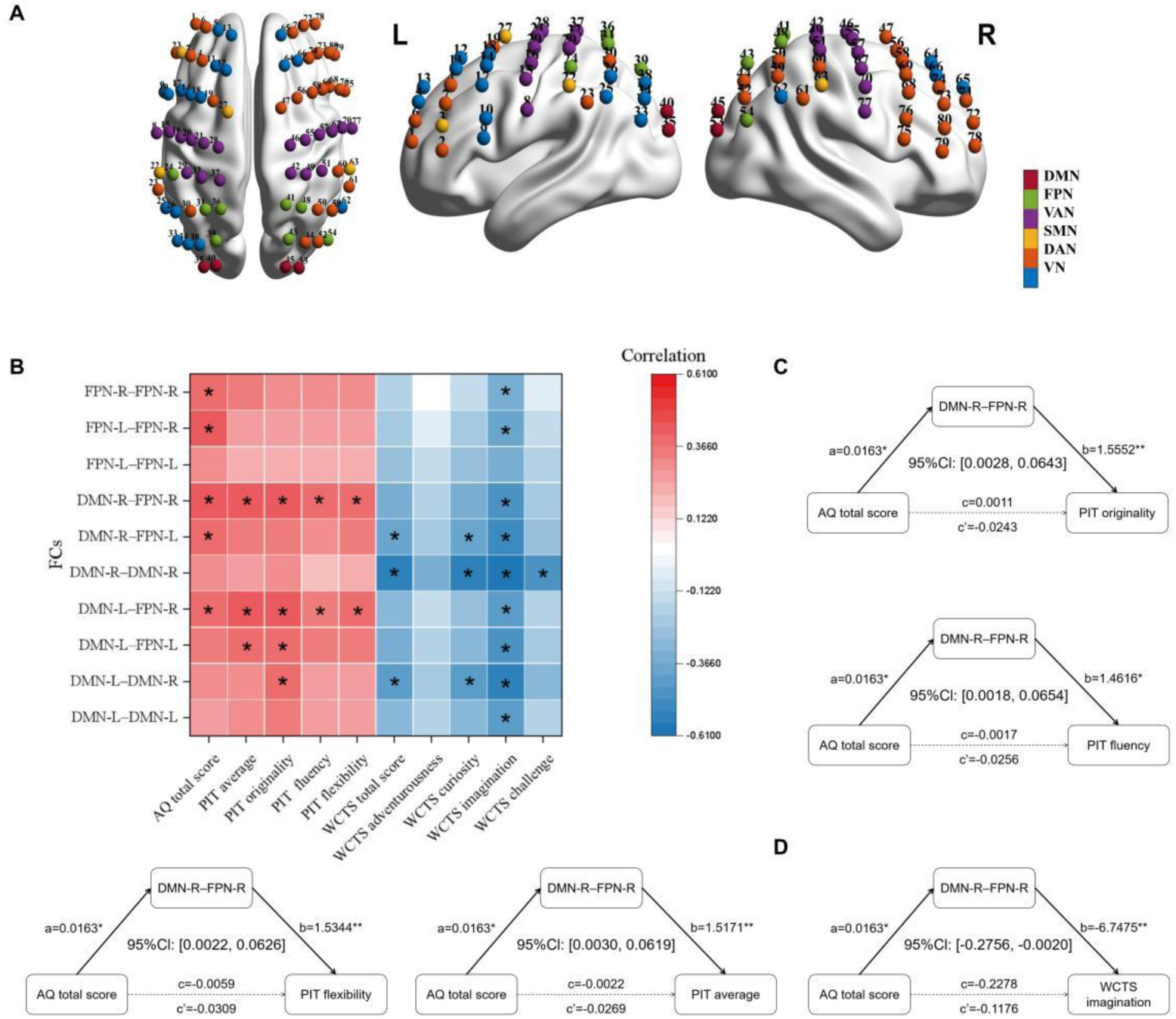
Correlation patterns and mediation models linking ATs, FCs, and creativity outcomes in ADHD. Note: (A) The arrangement of 80 measurement channels. (B) Heatmap shows partial correlations (controlling for age, sex and IQ) between FC indices and behavioral measures using listwise exclusion. Asterisks indicate *p* < 0.05 (uncorrected). (C-D) Mediation path models illustrate the indirect effects of ATs on PIT performance and WCTS imagination via DMN-R– FPN-R FC. Confidence intervals are bias-corrected bootstrap 95% CIs (5,000 resamples). \**p* < 0.05; \*\**p* < 0.01.

R_DMN-R_FPN significantly mediated the effect of ATs on PIT performance. ATs positively predicted R_DMN-R_FPN, which in turn positively predicted performance-based creativity indices from the PIT, including originality (*β* = 1.56, *p* = 0.007), fluency (*β* = 1.46, *p* = 0.018), flexibility (*β* = 1.53, *p* = 0.010), and the average score (*β* = 1.52, *p* = 0.007). The indirect effects of ATs on these indices through R_DMN-R_FPN were all significant (95% CI did not include zero), whereas neither the total nor direct effects were significant (**Figure 3C**).

In contrast, R_DMN-R_FPN significantly mediated the negative association between ATs and self-reported real-life creativity. Higher ATs were associated with stronger R_DMN-R_FPN, which in turn predicted lower scores on the “Imagination” subscale of the WCTS (*β* = −6.75, *p* = 0.029; 95% CI [−0.276, −0.002]) (**Figure 3D**).

### 3.4 Sensitivity analysis

To ensure the robustness of our findings, we conducted additional sensitivity analyses.

While earlier studies have suggested that intelligence and creativity are distinct constructs, to avoid the potential impact of intelligence on creativity necessitated the inclusion of years of education as a covariate in our regression models predicting creativity outcomes [43]. After controlling for years of education, the core findings remained largely unchanged. Group differences in DT, particularly across all PIT indices (originality, fluency, flexibility, and average scores), still remained statistically significant (all *p* <0.05). For real-life creativity, the group difference remained significant for the WCTS Imagination subscale (*p* =0.009). Importantly, years of education did not independently account for the observed advantages, suggesting that the creativity-related group differences were not solely driven by intelligence (see Supplementary **Table S2**).

In this study, 7 ADHD patients used medications. When controlling for medication status, the pattern of results remained unchanged.

## 4. Discussion

Building upon our integrative assessment of creativity and resting-state neural connectivity, this study examined how co-occurring autistic traits may contribute to heterogeneity in creativity among adults with ADHD. Our findings reveal that while ADHD at the group level is associated with heightened creative potential, elevated ATs were linked to a dimension-specific creativity profile: higher ATs were indirectly associated with enhanced DT performance and lower self-reported imagination via stronger right DMN–FPN functional connectivity. These findings provide preliminary evidence that right DMN– FPN connectivity may be involved in the divergent associations between ATs and different dimensions of creativity.

We found that ADHD outperformed controls on PIT specifically. However, no significant differences were found on the RAT, which is the most common test for CT. The dissociation between DT and CT further underscores the domain-specific nature of creative strengths in ADHD. These results are consistent with prior work [44, 45] and review [5]. In addition, we found individuals with ADHD exhibited generally stronger real-life creativity, as reflected in higher scores on the WCTS. This implies that these ideational strengths of ADHD may translate into real-life creative engagement and production. Further analyses showed that hyperactivity/impulsivity symptoms were more positively associated with creativity than inattentive symptoms. This pattern is consistent with a recent study showing that adults with the combined presentation of ADHD, characterized by pronounced HI symptoms, displayed higher creative potential than those with the inattentive presentation and controls [46]. Previous studies have linked ADHD-related distractibility and impulsivity to heightened curiosity and exploratory information-seeking, factors that may be conducive to divergent idea generation [47, 48]. Neurocognitive models also suggest that HI symptoms have a strong overlap with curiosity in terms of their neural substrates, such as the dopaminergic circuits involving the prefrontal cortex, which may enhance exploratory cognition and the generation of novel ideas [48, 49]. This may partly explain our results. In addition, it is interesting that our exploratory analyses revealed that impulsivity, rather than hyperactivity, was the primary driver of this association, suggesting that the creative advantage in ADHD may be more strongly linked to impulsive traits. This dissociation between impulsivity and hyperactivity may reflect their distinct cognitive underpinnings [50]. Speculatively, impulsivity-related reductions in inhibitory control or lower thresholds for response initiation may facilitate the generation of unconventional associations and ideas.

To explore how ATs influence creativity within the ADHD population, we divided participants into high (ADHD_AT+_) and low (ADHD_AT–_) autistic-traits subgroups. Results showed that the two subgroups exhibited distinct creative profiles when compared with the controls. Specifically, the ADHD_AT+_ group demonstrated a robust advantage in DT. This pattern indicates that ATs may be associated with certain originality-related aspects of DT within the ADHD population, consistent with prior evidence that autistic traits are linked to more unusual responses in divergent thinking tasks [18]. This may reflect a more persistent or detail-focused cognitive style associated with autistic traits, which could support creative performance during idea generation [51].

However, this DT advantage in the ADHD_AT+_ group did not translate into broadly enhanced real-life creativity, as no significant group differences were observed across most achievement-based measures. Consistent with thi<u>s</u>, partial correlation analyses revealed that AQ scores tended to be negatively correlated with several real-life creativity measures. Although these associations did not reach statistical significance, they suggest a possible dissociation between ideational potential and real-world creative accomplishments. Prior study has reported lower novelty seeking in autistic individuals [52]. One possible explanation is that elevated ATs may be associated with reduced novelty-seeking or risk-taking in uncertain real-world contexts, as autistic adults tend to avoid unpredictability in decision-making tasks [53]. In addition, reduced social motivation has been proposed as an important feature of autism [54]. This may limit opportunities for social engagement, collaboration, and feedback, thereby potentially hindering the translation of creative ideas into real-world outcomes. Together, these factors may help explain why ATs were linked to stronger DT but not to higher real-life creative expression.

We analyzed resting-state fNIRS data to identify potential neural mechanisms of creativity differences related to ATs within ADHD. Neuroscientific research of creativity has increasingly highlighted the involvement of large-scale brain networks in creative cognition, particularly the DMN and the FPN [36, 37]. Recent models of creative thinking propose that the dynamic interaction between these two networks facilitates the generation and evaluation of creative ideas: the DMN enables spontaneous ideation, whereas the FPN guides, constrains, and refines these ideas in accordance with task goals [36]. However, despite substantial progress in understanding these mechanisms in general populations, few studies have investigated the brain function of these creativity-related networks in individuals with ADHD. Even fewer studies have examined how these interactions may differ across ADHD subgroups such as those stratified by ATs.

Our mediation analysis provides evidence for a potential network-level pathway linking ATs to creativity in ADHD. Specifically, higher AQ scores predicted stronger DMN–FPN coupling, which in turn was associated with better performance-based DT (PIT originality, fluency, and flexibility), but lower scores in creative personality traits (e.g., WCTS imagination). This pattern highlights the role of right-lateralized DMN–FPN connectivity in creative cognition. Stronger DMN–FPN coupling may enhance the interplay between spontaneous ideation (DMN) and top-down control (FPN), facilitating divergent idea generation [36, 37]. This interpretation is broadly consistent with resting-state FC evidence linking divergent thinking and creative achievement to large-scale networks including the DMN and FPN-related systems [32]. However, the fact that the same DMN-FPN coupling showed an opposite mediated association with self-reported real-life creativity, highlights a dissociation between performance-based and self-reported aspects of creativity. We speculate that enhanced DMN–FPN connectivity primarily supports the generation stage of creative cognition, such as idea generation in DT tasks. However, it may not necessarily facilitate the subsequent stages of evaluation, refinement, and implementation that are essential for real-world creative accomplishment.

This study has several limitations that should be acknowledged. First, the limited size of the sample may limit the generalizability of results. However, our study is the first to explore how ATs relate to creativity in adults with ADHD, as well as the underlying brain mechanism. The inclusion of additional samples in future analyses would offer further insights. Second, control participants were recruited from the local community and did not undergo psychiatrist-administered diagnostic interviews; therefore, undetected neurodevelopmental or psychiatric traits cannot be fully excluded. Third, we used a cross-sectional design, which limits causal inferences about the relationships among ADHD, ATs, and creativity. Currently, the influence of ADHD and ATs on creative development over time remains insufficiently explored. Future research is advised to utilize longitudinal designs to trace causal relationships and determine whether the observed phenomena arise from innate factors, environmental influences, or compensatory adaptations. Finally, creativity is a multidimensional construct that can be assessed through various tasks. Although the present study utilized multiple methods to attempt a comprehensive assessment of creativity, it is possible that the full scope of creativity was not entirely captured. There is a need to develop and integrate more effective assessment approaches to provide a more thorough evaluation.

Through a study of rigorously diagnosed adults with ADHD and using a systematic creativity assessment battery, we have delineated the complex influence of ATs. We reveal that creativity in adults with ADHD is characterized by enhanced ideational divergence and real-world creative expression. ATs present a nuanced influence, subtly strengthening ideational divergence while potentially limiting practical creative expression. Right DMN–FPN connectivity may contribute to this discrepancy. Future interventions should focus on harnessing the generative cognitive style associated with ATs while circumventing their impediments to practical execution, thereby unlocking full creative potential and enhancing long-term outcomes.

## Acknowledgement

The authors would like to thank all participants who voluntarily took part in this study, as well as the research team of Peking University Sixth Hospital and Southwest University for their support in data collection and technical assistance.

## Authors’ contributions

Ningning Liu and Zelin Mu contributed equally to this work and share the first authorship. Jiang Qiu, Qiujin Qian, and Lu Liu are the corresponding authors, who had full access to all the data in the study. Gaoding Jia, Alitanshunbuer, and Jiabin Liu participated in data collection and preliminary data sorting. Xueyang Wang, Jinhua Xu, Haimei Li, and Yufeng Wang contributed to data analysis and manuscript revision. All authors reviewed, edited, and approved the final version of the manuscript.

## Ethical considerations

This study was approved by the Research Ethics Review Board (IRB) of Peking University Sixth Hospital (Approval Number: 2024-Ethics Rev.16) on March 19, 2024. All procedures were conducted in accordance with the ethical standards of the institutional research committee and with the 1964 Helsinki Declaration and its later amendments or comparable ethical standards. Written informed consent to participate was obtained from all participants prior to enrollment. As all participants were adults aged 18–45 years, parental consent was not applicable.

## Data availability

The datasets generated and/or analyzed during the current study are not publicly available due to the privacy of the participants. However, data are available from the authors upon reasonable request and with the permission of the participants.

## List of abbreviations

ADHD: Attention-deficit/hyperactivity disorder
AQ: Autism-Spectrum Quotient
ATs: Autistic Traits
DT: Divergent Thinking
CT: Convergent Thinking
AUT: Alternate Uses Task
PIT: Product Improvement Task
RAT: Remote Associates Task
CAQ: Creative Achievement Questionnaire
CBI: Creative Behavior Inventory
WCTS: Williams Creative Tendencies Scale
DSM-5: Diagnostic and Statistical Manual of Mental Disorders, Fifth Edition
DMN: Default Mode Network
FDR: False Discovery Rate
FC: Functional Connectivity
FPN: Frontoparietal Network
fNIRS: Functional Near-Infrared Spectroscopy
NT: Neurotypical controls
HI: Hyperactive/Impulsive (symptoms of ADHD)
IMP: Impulsivity (subscale of ADHD symptoms)
IQ: Intelligence Quotient
IA: Inattentive subtype (of ADHD)
MNI: Montreal Neurological Institute
NDDs: Neurodevelopmental Conditions

