## Supplementary Materials for "Accentuated Divergence, Constrained Expression: Autistic Traits Modulate the Creative Brain in Adults with ADHD"

### Supplementary Materials for Method

#### 1. Creative assessments

AUT: Responses were scored on three different outcome measures of DT: 1) fluency reflects the number of non-redundant responses, 2) flexibility is assessed by the number of conceptual categories the responses belong to, and 3) originality reflects how novel and uncommon the ideas are on a 5-point Likert scale (rated from 1 = not original at all, to 5 = very original). For each item, three trained coders counted all ideas to determine fluency and coded all ideas to determine flexibility and originality. Fleiss' Kappa coefficients for fluency, flexibility, and originality were 0.82, 0.76, and 0.79, respectively. For each dimension, scores from the "brick" and "can" items were first averaged across the two objects. Subsequently, these composite scores were standardized (z-transformed) across participants for further statistical analysis.

PIT: Participants were shown a schematic of a toy elephant and asked to list the most ingenious, interesting and unusual ways they can imagine to improve the toy and make it more enjoyable for children, without thinking about what will happen to the price. Responses were also scored on fluency, flexibility and originality of DT. For each item, three trained coders counted all ideas to determine fluency and coded all ideas to determine flexibility and originality. Fleiss' Kappa coefficients for fluency, flexibility, and originality were 0.75, 0.58, and 0.68, respectively.

CAQ: The CAQ evaluates creative achievement across ten domains: visual arts, music, dance, architectural design, creative writing, humor, inventions, scientific discovery, theater and film, and culinary arts. Each domain includes eight questions, which are ranked on a 0-7 scale: 0 = No training or recognized talent; 7 = National recognition of the participant's work. The total score is the sum of the scores across all domains.

WCTS: The WCTS consists of 50 items assessing trait-level creative tendencies. The questionnaire is scored on a 3-point Likert scale from 1 (totally disagree) to 3 (totally agree), with seven reverse-scored items. It is divided into four subscales: Curiosity (14 items); Imagination (13 items); Challenge (12 items); Adventurousness (11 items).

CBI: The CBI assesses everyday creativity across 28 items in literary, performing, visual arts, and crafts. Participants were asked to report how many times they engaged in each creative activity: 0 = Never; 1 = Once or twice; 2 = 3-5 times; 3 = More than five times. The total score is the sum of all individual scores.

**2. fNIRS Data Acquisition and Processing**

Data Acquisition: The fNIRS cap was fitted using the vertex and external auditory canals as anatomical reference landmarks following the international 10-20 positioning system. During the recording, participants sat in a quiet, dim room and and stay awake without movement.

Data Preprocessing: First, channels lacking a visible cardiac pulsation (~1 Hz) were excluded. On average 15.5 channels per participant were removed. Then the raw intensity signals were converted into optical density signals. Motion artifacts were corrected on a per-channel basis using spline interpolation within sliding 2-second windows. Signals were band-pass filtered between 0.01 and 0.1 Hz. Next, the relative hemoglobin concentration changes in oxygen-hemoglobin (HbO) and deoxygen-hemoglobin (HbR) were calculated via the modified Beer-Lambert Law. Finally, each participant's dataset was truncated to a stable 7-minute segment for further analysis.

FC calculation: , fNIRS channel positions were projected onto a standardized cortical surface based on the Yeo et al. network template, namely the default mode network (DMN), the frontoparietal network (FPN), the ventral attention network (VAN), the somatomotor network (SMN), the dorsal attention network (DAN) and the visual network (VN). The 10 DMN-FPN FC matrices generated for each participant are: DMN-R – DMN-R, DMN-R – DMN-L, DMN-R – FPN-R, DMN-R – FPN-L, DMN-L – DMN-L, DMN-L – FPN-R, DMN-L – FPN-L, FPN-R – FPN-R, FPN-R – FPN-L, FPN-L – FPN-L.

**Supplementary Materials for the Results**

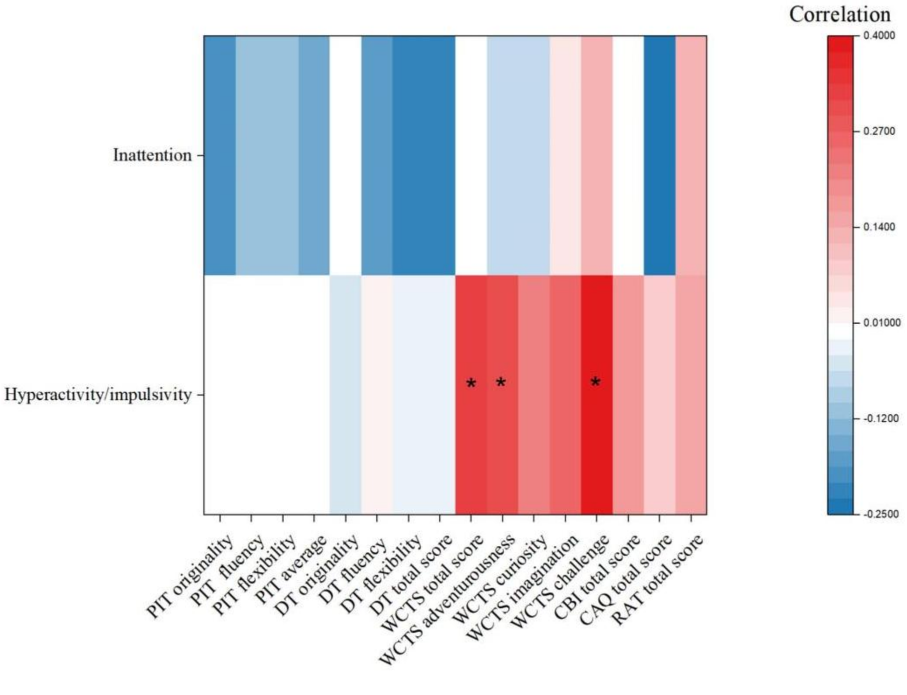


**Figure S1. Heatmap of the correlation between Hyperactivity/Inattention and creativity in ADHD**

Note：Significance is marked by the asterisk (*).

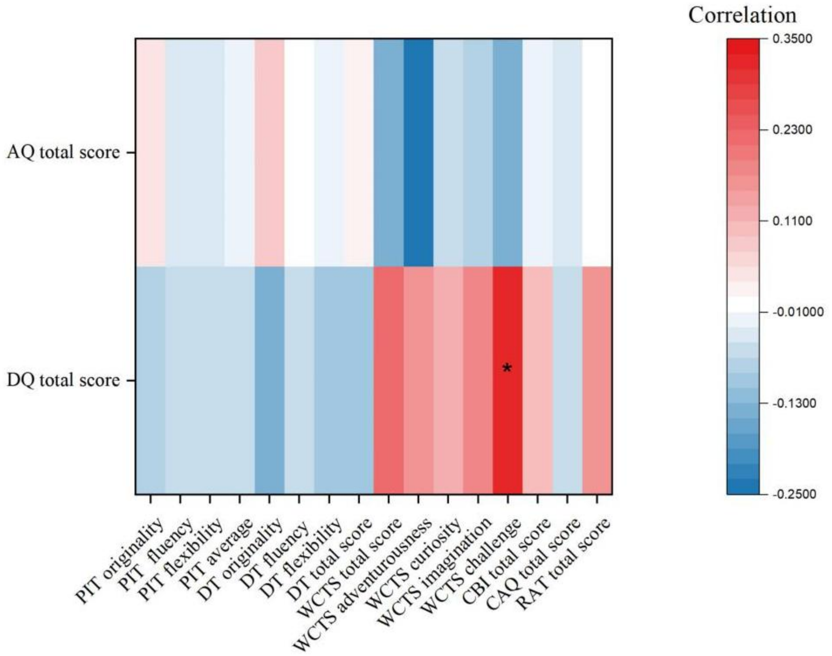


**Figure S2. Heatmap of the correlation between ASD and ADHD symptoms and creativity in ADHD**

Note：Significance is marked by the asterisk (*).

**Table S1. Multiple regression analyses exploring ASD and ADHD as predictors of creativity**

| Criterion predictor | Unstandardized Coefficients | | Standardized Coefficients | t | Sig. |
|---|---|---|---|---|---|
| | B | Std.error | Beta | | |
| **Divergent thinking** | | | | | |
| **PIT originality** | | | | | |
| **AQ total score** | 0.002 | 0.023 | 0.012 | 0.071 | 0.944 |
| **DQ total score** | -0.008 | 0.017 | -0.073 | -0.455 | 0.652 |
| **Sex** | -0.257 | 0.323 | -0.129 | -0.795 | 0.432 |
| **Age(years)** | 0.015 | 0.03 | 0.082 | 0.504 | 0.617 |
| **IQ** | 0.018 | 0.013 | 0.216 | 1.356 | 0.183 |
| **PIT fluency** | | | | | |
| **AQ total score** | -0.001 | 0.024 | -0.006 | -0.04 | 0.968 |
| **DQ total score** | -0.005 | 0.017 | -0.051 | -0.314 | 0.755 |
| **Sex** | 0.131 | 0.343 | 0.062 | 0.383 | 0.704 |
| **Age(years)** | -0.004 | 0.031 | -0.02 | -0.127 | 0.9 |
| **IQ** | 0.02 | 0.015 | 0.226 | 1.386 | 0.174 |
| **PIT flexibility** | | | | | |
| **AQ total score** | -0.061 | 0.061 | -0.166 | -0.992 | 0.328 |
| **DQ total score** | -0.002 | 0.024 | -0.01 | -0.064 | 0.95 |
| **Sex** | -0.006 | 0.017 | -0.051 | -0.32 | 0.751 |

| | | | | | |
|---|---|---|---|---|---|
| **Age(years)** | 0.128 | 0.34 | 0.06 | 0.375 | 0.71 |
| **IQ** | 0 | 0.031 | 0 | 0 | 1 |
| **PIT average** | | | | | |
| **AQ total score** | 0.002 | 0.023 | 0.015 | 0.098 | 0.923 |
| **DQ total score** | -0.007 | 0.017 | -0.066 | -0.407 | 0.686 |
| **Sex** | 0.114 | 0.333 | 0.056 | 0.344 | 0.733 |
| **Age(years)** | -0.005 | 0.03 | -0.025 | -0.154 | 0.879 |
| **IQ** | 0.02 | 0.014 | 0.23 | 1.409 | 0.167 |
| **DT originality** | | | | | |
| **AQ total score** | 0.005 | 0.02 | 0.04 | 0.26 | 0.796 |
| **DQ total score** | -0.014 | 0.015 | -0.146 | -0.933 | 0.357 |
| **Sex** | 0.08 | 0.291 | 0.043 | 0.276 | 0.784 |
| **Age(years)** | -0.036 | 0.026 | -0.214 | -1.373 | 0.178 |
| **IQ** | 0.019 | 0.013 | 0.234 | 1.489 | 0.145 |
| **DT fluency** | | | | | |
| **AQ total score** | -0.007 | 0.021 | -0.054 | -0.345 | 0.732 |
| **DQ total score** | -0.006 | 0.016 | -0.062 | -0.391 | 0.698 |
| **Sex** | 0.071 | 0.307 | 0.037 | 0.233 | 0.817 |
| **Age(years)** | -0.03 | 0.028 | -0.174 | -1.095 | 0.28 |
| **IQ** | 0.016 | 0.013 | 0.195 | 1.215 | 0.232 |
| **DT flexibility** | | | | | |
| **AQ total score** | -0.011 | 0.021 | -0.078 | -0.51 | 0.613 |
| **DQ total score** | -0.01 | 0.015 | -0.098 | -0.625 | 0.535 |
| **Sex** | 0.039 | 0.301 | 0.02 | 0.13 | 0.897 |
| **Age(years)** | -0.028 | 0.027 | -0.162 | -1.036 | 0.307 |
| **IQ** | 0.022 | 0.013 | 0.264 | 1.676 | 0.102 |
| **DT total** | | | | | |
| **AQ total score** | -0.013 | 0.061 | -0.032 | -0.209 | 0.835 |
| **DQ total score** | -0.03 | 0.045 | -0.105 | -0.666 | 0.509 |
| **Sex** | 0.189 | 0.878 | 0.034 | 0.216 | 0.83 |
| **Age(years)** | -0.095 | 0.079 | -0.187 | -1.195 | 0.239 |
| **IQ** | 0.057 | 0.038 | 0.238 | 1.501 | 0.141 |
| **Creative Achievement** | | | | | |
| **WCTS total score** | | | | | |
| **AQ total score** | -0.259 | 0.258 | -0.128 | -1.002 | 0.321 |
| **DQ total score** | 0.316 | 0.188 | 0.219 | 1.682 | 0.099 |
| **Sex** | -0.56 | 3.704 | -0.02 | -0.151 | 0.88 |
| **Age(years)** | -0.111 | 0.335 | -0.043 | -0.331 | 0.742 |
| **IQ** | 0.42 | 0.16 | 0.346 | 2.633 | 0.011* |
| **WCTS adventurousness** | | | | | |
| **AQ total score** | -0.105 | 0.064 | -0.221 | -1.651 | 0.105 |
| **DQ total score** | 0.063 | 0.046 | 0.188 | 1.37 | 0.177 |
| **Sex** | -0.181 | 0.912 | -0.027 | -0.198 | 0.844 |
| **Age(years)** | 0.011 | 0.082 | 0.019 | 0.137 | 0.891 |

| | | | | | |
|---|---|---|---|---|---|
| IQ | 0.051 | 0.039 | 0.178 | 1.287 | 0.204 |
| **WCTS curiosity** | | | | | |
| AQ total score | -0.032 | 0.089 | -0.046 | -0.359 | 0.721 |
| DQ total score | 0.056 | 0.065 | 0.114 | 0.859 | 0.394 |
| Sex | -0.495 | 1.274 | -0.051 | -0.389 | 0.699 |
| Age(years) | -0.08 | 0.115 | -0.092 | -0.696 | 0.489 |
| IQ | 0.16 | 0.055 | 0.388 | 2.91 | 0.005* |
| **WCTS imagination** | | | | | |
| AQ total score | -0.061 | 0.09 | -0.087 | -0.674 | 0.504 |
| DQ total score | 0.092 | 0.066 | 0.186 | 1.401 | 0.167 |
| Sex | 0.047 | 1.298 | 0.005 | 0.036 | 0.971 |
| Age(years) | -0.074 | 0.117 | -0.083 | -0.627 | 0.534 |
| IQ | 0.134 | 0.056 | 0.321 | 2.4 | 0.02* |
| **WCTS challenge** | | | | | |
| AQ total score | -0.062 | 0.065 | -0.121 | -0.967 | 0.338 |
| DQ total score | 0.119 | 0.047 | 0.326 | 2.538 | **0.014*** |
| Sex | -0.021 | 0.926 | -0.003 | -0.023 | 0.982 |
| Age(years) | 0.034 | 0.084 | 0.053 | 0.41 | 0.683 |
| IQ | 0.092 | 0.04 | 0.299 | 2.314 | 0.025* |
| **CBI total score** | | | | | |
| AQ total score | -0.135 | 0.267 | -0.069 | -0.506 | 0.615 |
| DQ total score | 0.149 | 0.194 | 0.107 | 0.766 | 0.447 |
| Sex | 6.112 | 3.827 | 0.223 | 1.597 | 0.116 |
| Age(years) | -0.093 | 0.346 | -0.037 | -0.268 | 0.79 |
| IQ | -0.082 | 0.165 | -0.07 | -0.498 | 0.621 |
| **CAQ total score** | | | | | |
| AQ total score | -0.144 | 0.272 | -0.075 | -0.53 | 0.598 |
| DQ total score | -0.069 | 0.198 | -0.05 | -0.346 | 0.731 |
| Sex | 1.307 | 3.901 | 0.048 | 0.335 | 0.739 |
| Age(years) | -0.003 | 0.353 | -0.001 | -0.009 | 0.993 |
| IQ | 0.074 | 0.168 | 0.064 | 0.438 | 0.663 |

Note： AQ: Autism - Spectrum Quotient; CAQ: Creative Achievement Questionnaire; CBI: Creative Behavior Inventory; DQ: ADHD-RS-IV total score DT: Divergent thinking; IQ: Intelligence Quotient; PIT: Product Improvement Task; WCTS: Williams Creative Tendencies Scale; *p<0.05, **p<0.01.

**Table S2. Multiple regression analyses predicting divergent thinking indices with group (ADHD vs. NT) and demographic covariates**

| Dependent Variable | Unstandardized Coefficients | | Standardized Coefficients | t | Sig. |
|---|---|---|---|---|---|
| | B | Std.error | Beta | | |

| | | | | | |
|---|---|---|---|---|---|
| **Divergent thinking** | | | | | |
| **PIT originality** | | | | | |
| **Group** | -0.59 | 0.23 | -0.261 | -2.563 | **0.012*** |
| **Sex** | 0.149 | 0.249 | 0.057 | 0.599 | 0.55 |
| **Age** | 0.019 | 0.022 | 0.086 | 0.89 | 0.375 |
| **Year of Education** | 0.115 | 0.146 | 0.083 | 0.79 | 0.431 |
| **PIT fluency** | | | | | |
| **Group** | -3.092 | 1.489 | -0.215 | -2.076 | **0.04*** |
| **Sex** | 0.393 | 1.61 | 0.023 | 0.244 | 0.808 |
| **Age** | -0.003 | 0.141 | -0.002 | -0.021 | 0.983 |
| **Year of Education** | 1.283 | 0.945 | 0.145 | 1.358 | 0.177 |
| **PIT flexibility** | | | | | |
| **Group** | -2.723 | 1.313 | -0.214 | -2.074 | **0.04*** |
| **Sex** | 0.331 | 1.42 | 0.022 | 0.233 | 0.816 |
| **Age** | 0.019 | 0.125 | 0.015 | 0.155 | 0.877 |
| **Year of Education** | 1.213 | 0.833 | 0.155 | 1.456 | 0.148 |
| **PIT average** | | | | | |
| **Group** | -6.405 | 2.993 | -0.221 | -2.14 | **0.035*** |
| **Sex** | 0.873 | 3.237 | 0.026 | 0.27 | 0.788 |
| **Age** | 0.036 | 0.284 | 0.012 | 0.126 | 0.9 |
| **Year of Education** | 2.611 | 1.899 | 0.146 | 1.375 | 0.172 |
| **DT originality** | | | | | |
| **Group** | -0.209 | 0.208 | -0.102 | -1.005 | 0.317 |
| **Sex** | 0.245 | 0.227 | 0.104 | 1.079 | 0.283 |
| **Age** | -0.006 | 0.019 | -0.033 | -0.334 | 0.739 |
| **Year of Education** | -0.063 | 0.138 | -0.047 | -0.458 | 0.648 |
| **DT fluency** | | | | | |
| **Group** | 0.083 | 0.672 | 0.013 | 0.123 | 0.902 |
| **Sex** | 0.771 | 0.735 | 0.102 | 1.049 | 0.296 |
| **Age** | -0.048 | 0.063 | -0.076 | -0.768 | 0.444 |
| **Year of Education** | -0.046 | 0.446 | -0.011 | -0.102 | 0.919 |
| **DT flexibility** | | | | | |
| **Group** | 0.203 | 0.6 | 0.035 | 0.339 | 0.735 |
| **Sex** | 0.81 | 0.655 | 0.12 | 1.236 | 0.219 |
| **Age** | -0.033 | 0.056 | -0.058 | -0.593 | 0.554 |
| **Year of Education** | 0.158 | 0.398 | 0.041 | 0.398 | 0.692 |
| **DT total** | | | | | |
| **Group** | 0.077 | 1.444 | 0.005 | 0.053 | 0.958 |
| **Sex** | 1.826 | 1.578 | 0.112 | 1.158 | 0.25 |
| **Age** | -0.088 | 0.134 | -0.064 | -0.652 | 0.516 |
| **Year of Education** | 0.049 | 0.958 | 0.005 | 0.052 | 0.959 |
| **Creative Achievement** | | | | | |
| **WCTS total score** | | | | | |
| **Group** | -3.614 | 2.895 | -0.124 | -1.248 | 0.214 |

| | | | | | |
|---|---|---|---|---|---|
| **Sex** | -1.105 | 3.065 | -0.033 | -0.361 | 0.719 |
| **Age** | -0.309 | 0.268 | -0.107 | -1.151 | 0.252 |
| **Year of Education** | 0.451 | 1.929 | 0.024 | 0.234 | 0.815 |
| **WCTS adventurousness** | | | | | |
| **Group** | 0.122 | 0.65 | 0.019 | 0.187 | 0.852 |
| **Sex** | -0.86 | 0.688 | -0.116 | -1.25 | 0.214 |
| **Age** | -0.021 | 0.06 | -0.033 | -0.349 | 0.728 |
| **Year of Education** | 0.079 | 0.433 | 0.019 | 0.183 | 0.855 |
| **WCTS curiosity** | | | | | |
| **Group** | -0.318 | 0.968 | -0.033 | -0.328 | 0.743 |
| **Sex** | -0.696 | 1.025 | -0.063 | -0.68 | 0.498 |
| **Age** | -0.116 | 0.09 | -0.122 | -1.297 | 0.197 |
| **Year of Education** | 0.381 | 0.645 | 0.06 | 0.591 | 0.555 |
| **WCTS imagination** | | | | | |
| **Group** | -2.619 | 0.983 | -0.257 | -2.664 | **0.009**** |
| **Sex** | 0.387 | 1.041 | 0.033 | 0.372 | 0.711 |
| **Age** | -0.114 | 0.091 | -0.113 | -1.248 | 0.214 |
| **Year of Education** | -0.079 | 0.655 | -0.012 | -0.121 | 0.904 |
| **WCTS challenge** | | | | | |
| **Group** | -0.799 | 0.676 | -0.117 | -1.181 | 0.24 |
| **Sex** | 0.064 | 0.716 | 0.008 | 0.09 | 0.929 |
| **Age** | -0.058 | 0.063 | -0.086 | -0.921 | 0.359 |
| **Year of Education** | 0.07 | 0.451 | 0.016 | 0.155 | 0.877 |
| **CBI total score** | | | | | |
| **Group** | -2.489 | 2.166 | -0.107 | -1.149 | 0.253 |
| **Sex** | 7.464 | 2.335 | 0.281 | 3.196 | 0.002** |
| **Age** | -0.23 | 0.204 | -0.101 | -1.128 | 0.262 |
| **Year of Education** | -1.531 | 1.439 | -0.102 | -1.064 | 0.289 |
| **CAQ total score** | | | | | |
| **Group** | 0.469 | 2.975 | 0.016 | 0.157 | 0.875 |
| **Sex** | -2.362 | 3.15 | -0.069 | -0.75 | 0.455 |
| **Age** | -0.386 | 0.276 | -0.13 | -1.4 | 0.164 |
| **Year of Education** | 2.901 | 1.983 | 0.147 | 1.463 | 0.146 |

Note：Variable coding: Group: 1 = ADHD group, 2 = neurotypical (NT) group; Sex: 1 = male, 2 = female. AQ: Autism - Spectrum Quotient; CAQ: Creative Achievement Questionnaire; CBI: Creative Behavior Inventory; DQ: ADHD-RS-IV total score ; DT: Divergent thinking; IQ: Intelligence Quotient; PIT: Product Improvement Task; WCTS: Williams Creative Tendencies Scale; *p<0.05, **p<0.01.
